# Tumor size>5cm is a line of demarcation of mortality and progression of gastric cancer after D2+gastrectomy for Chinese population

**DOI:** 10.1101/2023.12.10.23299776

**Authors:** Yifan Li, Hanliang Zhao, Yinan Shi, Ruirui Yang

## Abstract

**Methods:** Gastric cancer between May 2002 and December 2020 and who had undergone resection of the primary cancer. We analyzed these patients to study the association between survival and tumor size by Cox proportional hazards model and restricted cubic splines.

**Results:** A total of 1708 patients met the inclusion criteria, with a median age of 58 years. The distribution of tumor size was correlated with patients underwent different D2+ gastrectomy (P<0.001) and located different tumor site(P=0.002). The size of the patient’s tumor is closely related to the patient’s prognosis, as well as the overall survival of patients experienced proximal gastrectomy (P for trend= 0.002) and progression free survival of distal(P for trend= 0.03) and total gastrectomy (P for trend= 0.016) in fully adjusted model. Likewise, tumor size displayed its prognostic predictability in subgroup of upper 1/3, but only for overall survival in final model(P for trend= 0.045). Nonlinear relationship of different tumor size and D2+ gastrectomy or tumor site showed in restricted cubic splines, >5cm showed a significant impact in each group, but not for proximal gastrectomy (P for nonlinear=0.305). Overall survival and progression decreased progressively along with upgrading of tumor size accordingly.

**Conclusions:** Tumor size>5cm can be seen as a line of demarcation of mortality and progression of gastric cancer after D2+gastrectomy, the hazard ratio began to rise when tumor size large than 5cm.

## Background

One of the most common malignant tumors in the world is gastric cancer. Gastric cancer mainly occurs in the northwest and eastern coastal areas in my country ^[1–3]^. Today, around the world, the third of the junction between the human stomach and the stomach and esophagus has always been a high-risk area for cancer, and the incidence rate has been increasing year by year ^[4–5]^.

In order to manage cancer patients (including cancer) more effectively, the American Joint Commission on Cancer (AJCC) has proposed a cancer risk stratification system, which includes the tumor (T), lymph node (N), and metastasis (M) systems. Nowadays, this system for stratifying cancer risk has been widely recognized and applied in the medical community^[6]^. The allocation of clinical staging (cTNM) is based on radiographic and endoscopic evaluation, while pathological staging (pTNM) needs to be evaluated using excised specimens^[7]^. For patients with rectal cancer or primary gastric tumors who have received systemic or radiation therapy before cancer resection, these patients will be further classified. They are classified as neoadjuvant pathological stage (ypTNM) ^[8]^. In TNM staging, the prognostic significance of solid tumor size or horizontal growth can have significant implications for tumor prognosis, tumor recurrence, patient survival, and clinical management^[9]^. While spreading horizontally, cancer also penetrates deeper into various layers through the gastric wall. At present, the American Joint Commission on Cancer (AJCC) uses the degree of tumor infiltration as theT-stage standard for gastric cancer, rather than measuring it based on the size or horizontal spread of the tumor.

According to other retrospective studies, we can know that the size of the tumor or its maximum horizontal diameter will lead to the low survival rate of cancer patients ^[10]^. Although the sample size of most studies is very limited, additional research and analysis are still needed to use tumor size to estimate cancer survival. The research involved in this paper is based on the largest cohort study to date to assess the extent to which cancer survival is affected by the size and extent of gastric tumors.

## Methods

### Data collection ^[18]^

In the study, the researchers studied 1,708 patients who underwent surgery for gastric cancer between May 2002 and December 2020. All these patients underwent radical gastric surgery. Patients included in the study met the following requirements, one of which was that curative surgery for gastric cancer (R0-R1) and preliminary histological diagnosis had been completed. ^[18]^.

Study inclusion criteria: 1. The cancer has been confirmed by histopathology; 2. Total gastrectomy has been completed in patients with stage I, II, III, and IV cancer; 3. Complete clinical pathology and follow-up data can be provided (within 1 week before operation) Complete the measurement of all biomarkers required for the study); 4. After surgery, the patient did not show serious organ damage; 5. The patient had no other malignant tumors.

Study exclusion criteria: 1. Patients with other systemic tumors; 2. Patients unable to provide complete clinical data; 3. Patients with bypass surgery or palliative surgery; 4. Patients whose tumors were confirmed to be non-gastric cancer after pathological classification. This study was a retrospective analysis of patients with gastric cancer, so informed consent from patients could not be obtained. All procedures in this research process conform to the standards of the Declaration of Helsinki and have been reviewed and approved by the Ethics Committee of the Cancer Hospital. For patient data, the principles of anonymity and confidentiality are adopted^[18]^.

In addition to the above criteria, it also includes age, sex, type of gastrectomy, maximum tumor diameter, nerve invasion, vascular invasion, surgical margin, omentum metastasis, number of positive lymph nodes, TNM staging (according to the 8th edition of the United States Joint Commission), pT staging, Lauren classification, AE1/AE3, CD56, CDX-2, CGA, CK7, CK20, SATB-2, SYN, PMS2, MLH1, MSH2 MSH6, as well as progression free survival (PFS) and overall survival (OS). Follow-up time was determined from electronic records of patients’ visits with oncologists and hospital visit records. Follow-up time for patients was determined and calculated based on the date of the patient’s last visit, as well as the date of the last contact with the surgeon. The period from the time of surgery to the first discovery of tumor metastasis in the patient is called progression-free survival (PFS). The period from the completion of the operation to the death of the patient is called the overall survival (OS), which can also be calculated based on the last follow-up records of the patient ^[18]^. The study patients in this project were followed up until December 2020, and the follow-up time was mostly about 41.51 ± 21.18 months. Patients need to be followed up every 3 months within the first 1 year after surgery. Within 2 to 5 years after surgery, patients need to complete a follow-up visit every 6 months. After that, it is enough to complete the follow-up of patients once a year. Routine follow-up items include the following: patient physical examination, patient chest X-ray, pelvic ultrasound, magnetic resonance imaging and computed tomography, and laboratory tests..

### Statistical analysis

Tumor size is defined as maximum diameter of tumor and the greatest length, width or height in the boundary of tumor was seen as the actual size of the tumor. The division of tumor location as depicted in Figure 7, tumor located in cardia or fundus was upper 1/3, body of stomach was middle 1/3, antrum or pylorus was lower 1/3. According to the size of the tumor, it can be divided into three groups: tumors smaller than 2cm, tumors between 2-4cm, tumors between 4-6 cm, tumors between 6-8 cm, and tumors larger than 8cm. Summarize the pathological and clinical characteristics of the study population, as well as demographic characteristics, and describe the types and distribution of statistical variables. By analyzing the univariate and multivariate data of the study subjects, determine which factors are closely related to patient outcomes. All variables with clinical significance should be included for analysis and research. Afterwards, the significance levels of each of these variables were eliminated one by one. The association between patient survival rate and various indications was evaluated by fitting within the Cox proportional risk model using the forward elimination method (i.e. removal standard p=0.05). At a level of 0.05, remove the covariates with a p-value of 0.05 from the multivariate model. Then, through multivariate COX regression analysis, record the relevant factors between PFS and OS and analyze them. To explore the non-linear effects of overall survival and progression free survival on tumor size, the restricted cubic splines with six knots were introduced in the fully adjusted model using continuous measures of tumor size. Use relevant software including but not limited to SPSS 25.0, R software (version 4.1.2), and Grand pad Prism 9.3 to process the data.

## Results

### Patient Demographics

A total of 1708 patients met the eligibility criteria, with an average age of diagnosis of 58.75 ± 10.04 years. Gender distribution revealed male was 79.5% and male occupy a predominant number in each stage. The proportion of tumor location of the patients were 50.7% for upper 1/3, 16.5% for middle 1/3, 32.0% for lower 1/3, 0.8% for multiple, respectively. The ratio of older patients (>60 years old) and younger (<60 years old) was almost near, 52.8% and 47.2%, respectively. Distal gastrectomy was second highest number of D2+ radical gastrectomy, only next to total gastrectomy. Intestinal type (39.2%), diffuse type(35.6%) and mixed type(25.2%) nearly have equal shares in Lauren classification (Table 1).

### Impact of Tumor Size on Clinical Presentation and Tumor Characteristics

The distributions across stages I-IV were 19.67%, 24.12%, 52.81% and 3.4%; respectively. The primary tumors were almost less than 2-4cm in size (32.9%), followed by 4-6cm (30.9%), 6-8cm (15.3%), 0-2cm (12.1%) and >8cm (8.8%). Most of the tumors were adenocarcinoma (68.7%) and had positive expression of MLH1 (87.2%), PMS2(72.7%), MSH2(86.4%) and MSH6 (87.2%). Compared to tumors smaller than 2 centimeters in size, primary tumors larger than 8 centimeters are more likely to lead to the occurrence of advanced diseases (stage IV: 15.5% vs. 1.7%; p<0.001), and were less likely to be located lower 1/3 (16.1% vs. 4.4%; p<0.001). When undergoing radical resection, the probability of positive edges appearing on them is higher (22.0% vs. 6.1%; p<0.001), higher frequency of vascular invasion (10.7% vs. 2.7%; p<0.001), neural invasion (11.7% vs. 3.2%; p<0.001), more positive expression of AE1/AE3 (10.7% vs. 5.2%; p<0.001) and CD56 (10.7% vs. 4.7%; p<0.001). Most tumor size 2-4cm in upper1/3(41.0%) and lower 1/3(41.5%) received proximal gastrectomy and distal gastrectomy, respectively. By contrast, the great majority total gastrectomy applied for tumor size 2-4cm (26.8%), 4-6cm (33.1%), 4-6cm (21.6%) and >8cm(13.1%). Intestinal type occurred in tumor size<6cm(0-2cm:23.1%, 2-4cm:36.5%, 4-6cm:25.5%) compared to tumor size 2-4cm(26.0%), 4-6cm(34.0%) and>8cm(22.0%) for diffuse type. Primary tumor size<2cm have high level deficient mismatch repair(dMMR) (68.9% vs. 36.1% vs. 27.8% vs. 24.0% vs. 24.7%, p<0.001). (Table 2).

According to the stratification results of AJCC staging, 50.0% of patients with stage I tumors have tumor sizes ranging from 0 to 2 centimeters, 40.2% have tumor sizes ranging from 2 to 4 centimeters, and about 0.9% have tumors larger than 8 centimeters in their bodies (p<0.001). For stage IV cancer patients, only 1.7% had tumors smaller than 2 centimeters in size, and 74.2% had tumors larger than 4 centimeters in body (compared to 9.8% for stage I; p<0.001). 336 patients with stage I (pT1N1:80.9%; pT1N1:5.7%; pT2N0:13.4%) and 58 patients with stage IV cancer. Stage IV cancer (62.1%) is more than half likely to occur in patients under the age of 60; There is an 8.6% chance that it will occur in patients aged 70 or above; The probability of phase I occurring in patients under 60 years old and those over 70 years old is around 52.8%. Tumor located in upper 1/3 were more likely to across over stage I (41.7%) and stage IV (50.0%, P=0.439) and nearly half of lower 1/3(44.0%) were inclined to revealed in stage I by contrast with stage IV (29.3%, P=0.148). Similarly, almost 50 percent patients with stage I (44.8%) underwent distal gastrectomy (compared to 22.4% in stage IV; p<0.001) and 72.4% of stage IV received total gastrectomy (compared to 28.9% in stageI; p<0.001). Compared with 96.6% of patients with stage IV tumors (P=0.525), most patients undergoing stage I surgery showed a negative resection rate (97.9%). And patients with stage I cancer are less likely to experience omental metastasis in the later stage (0.3%) compared to stages IV (6.9%, P=0.004), more frequent presented intestinal type (81.5%) in comparison with stages IV (27.6%, P<0.001), and higher frequency of dMMR(stage I:71.7% vs. stage IV: 17.2%, P<0.001), and less regularly occurred diffuse type (stage I:7.1% vs. stage IV: 53.4%, P<0.001).

The study subjects included 412 patients with stage II cancer diseases (pT1-4, N0-3). The tumor size of patients is mostly between 2-4cm (42.0%) and 4-6 cm (34.7%). The number of patients with tumor sizes<2cm (5.6%) or>8cm (4.9%) is relatively small. More vascular invasion(40.5%), neural invasion(38.1%), total gastrectomy (58.3%), expression of AE1/AE3(79.1%), Her-2(41.5%), CD56(46.5%), CDX-2(46.6%) get involved in this stage. Most of tumor located at upper 1/3(88.6%) received total gastrectomy compared with less of tumor (11.4%) underwent proximal gastrectomy. Likewise, 92.1% tumor located at lower 1/3 experienced distal gastrectomy and others (7.9%) underwent total gastrectomy. The confirmed number of patients with stage III disease (Any T, N+) is 902. The tumor size distribution is: <2cm (1.6%), 2-4cm (26.6%), 4-6cm (37.4%), 6-8cm (21.4%)and >8cm (13.1%). Compared to stage I patients, there was a higher incidence of vascular invasion(75.5%), neural invasion(66.1%), total gastrectomy (67.3%), positive surgical margin(7.2%), omentum metastasis (4.7%), expression of AE1/AE3(94.3%) and CDX-2(49.1%). Age between 60 and 69 were more advantaged for stage II (40.5%) and III(35.8%) in terms of number of patients. Most of tumor located at upper 1/3(89.1%) received total gastrectomy compared with less of tumor (10.9%) underwent proximal gastrectomy. Likewise, 86.7% tumor located at lower 1/3 experienced distal gastrectomy and others (13.3%) underwent total gastrectomy.

We selected 58 GC patients. These patients’ cancers belong to stage IV and have undergone surgical resection of the primary tumor, which has undergone pathological AJCC T staging. Among them, 62.9% of patients are under the age of 60. The distribution of primary tumor size was <2cm (1.7%), 2-4cm (24.1%), 4-6cm (39.7%), 6-8cm (19.0%) and >8cm (15.5%). The proportion of cancer patients with tumor size<4cm (59.1%) was (p<0.001), while 4.4% of cancer patients with tumor size>8cm were in the lower one-third. If compared to patients with tumor size<4cm (6.1%; p<0.001), patients with tumor size>8cm are more likely to show positive margins after tumor resection (22.0% of patients show positive margins). Many tumor located at upper 1/3(89.6%) received total gastrectomy compared with less of tumor (10.4%) underwent proximal gastrectomy. Likewise, 76.5% tumor located at lower 1/3 experienced distal gastrectomy and others (23.5%) underwent total gastrectomy.

### Impact of Tumor Size with different gastrectomy on Survival

The median follow-up time of the cohort was 38.9 months, with no patients lost to follow-up. A total 1706 patients received radical gastrectomy enrolled in the first section of research(Figure 1).There was a significant association between tumor size and a increased risk of mortality for patients who accepted proximal gastrectomy after controlling for age, sex, pT stage number of positive lymph nodes (Adjusted^a^ model). Even in the further-adjusted model (Adjusted^b^ model), the harmful effect remained significant. The mortality risk ascended as tumor size upgraded in crude model(P for trend 0.013), Adjusted^a^ model(P for trend 0.017) or Adjusted^b^ model(P for trend 0.002). However, for patients who received total (P for trend 0.102) or distal gastrectomy (P for trend 0.112), the mortality risk between different tumor size seemed to be similar. The exploratory model showed similar results of subgroups of proximal (P for trend 0.002), total (P for trend 0.132) and distal (P for trend 0.12) gastrectomy. Nevertheless, the outcomes of Cox regression analysis for progression free survival(table 4) were exactly opposite. The difference of progression hazard for various tumor size was significant for patients who underwent total (P for trend =0.011) and distal gastrectomy (P for trend= 0.029) and the pernicious impact was more evidently in fully adjusted model. Interestingly enough, the adverse effect of tumor size was inconsistent in crude model(P for trend= 0.259), adjusted^a^ model(P for trend= 0.237) and final adjusted model (P for trend= 0.021) of proximal gastrectomy cohort. Meanwhile, we noticed that hazard ratio was not accrued as tumor size became larger.

**Figure 1.**
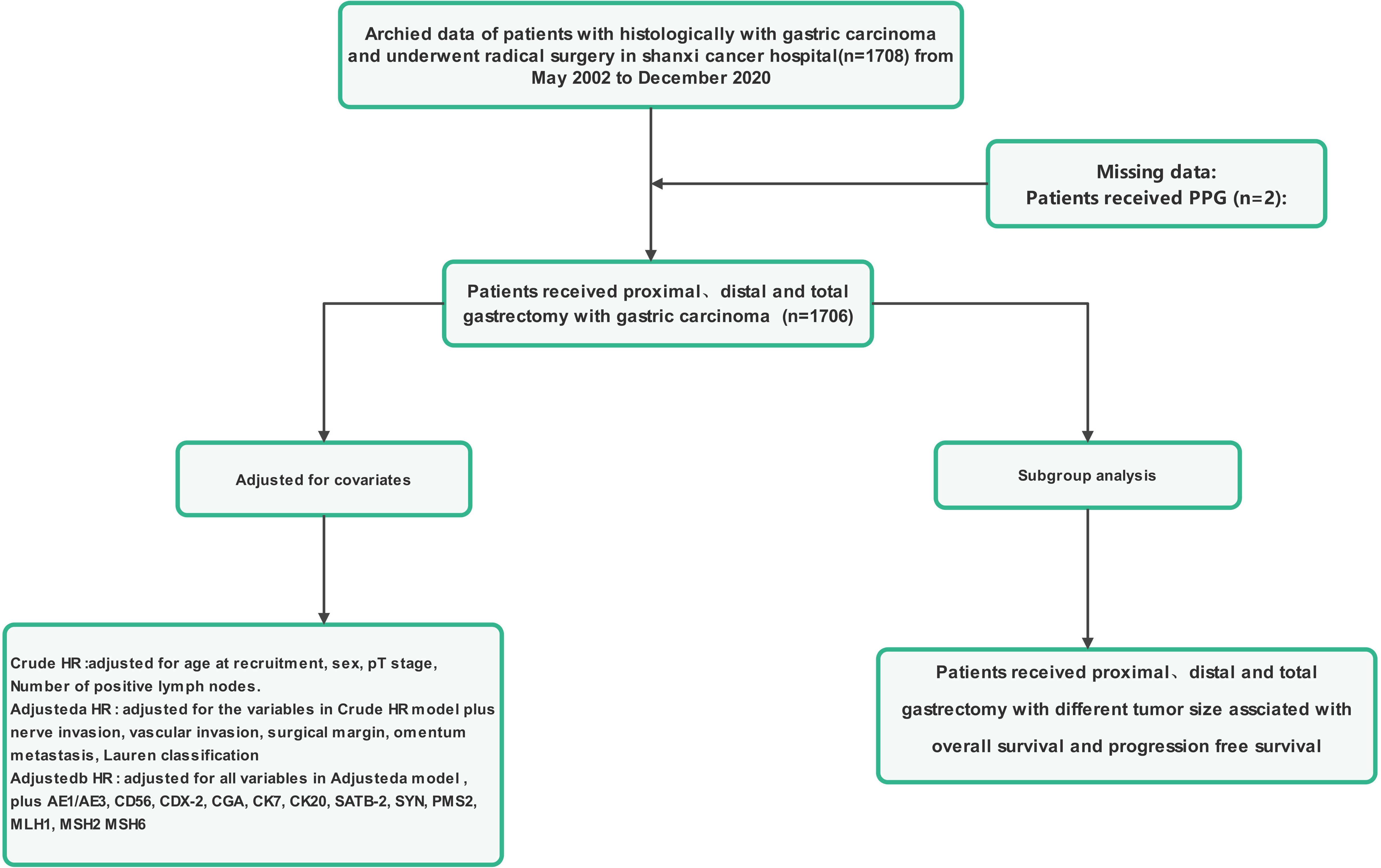
Flowchart for the enrollment of patients received D2+gastrectomy +.

**Figure 2.**
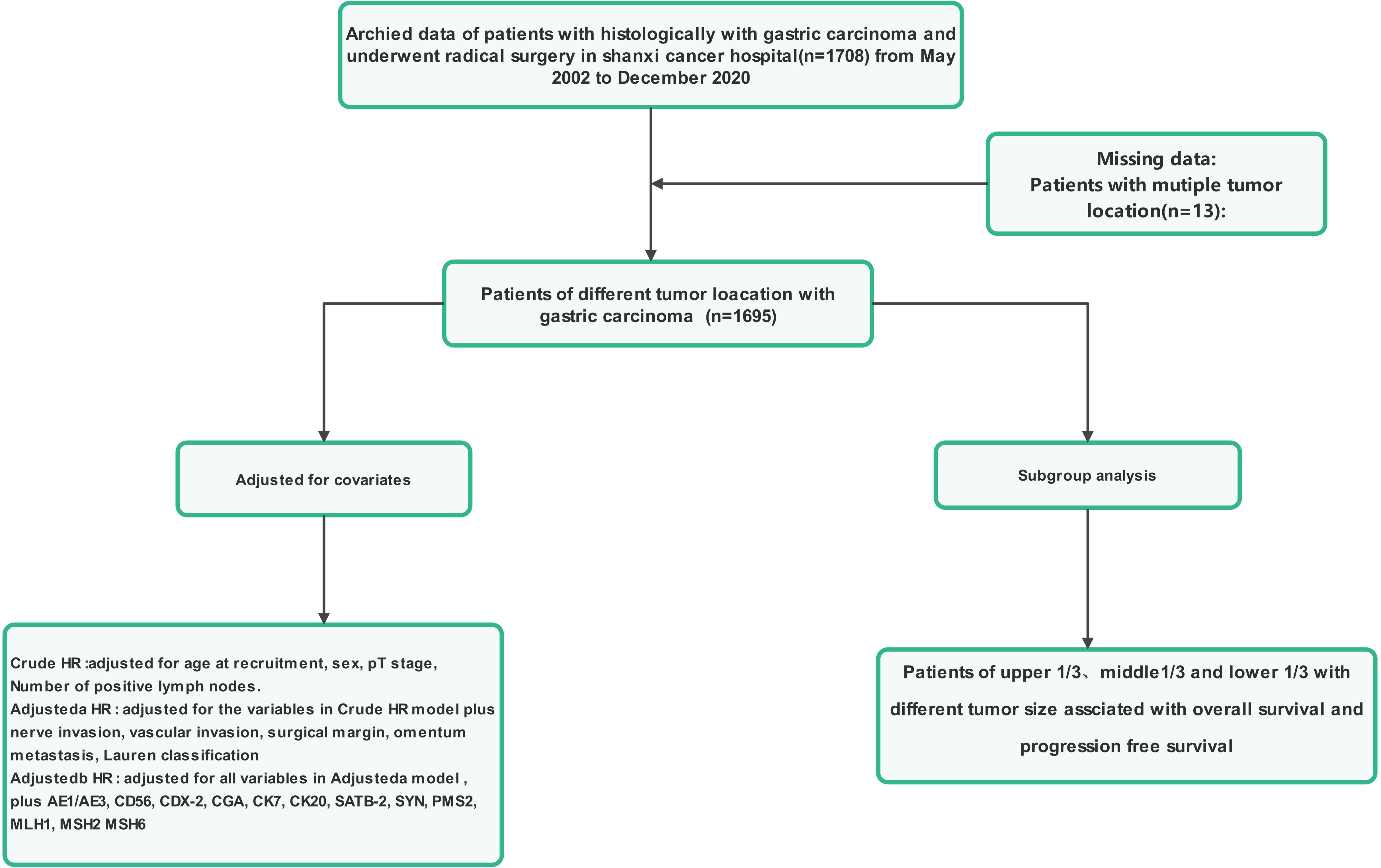
Flowchart for the enrollment of patients with different tumor location.

In Figure 3, we used restricted cubic splines(RCS) to flexibly model and visualize the association of tumor size with overall survival for patients experienced various gastrectomy. After adjusting for all the covariates, mortality risk steeply when tumor size more than 5cm for the whole cohort(P for nonlinear<0.001), distal gastrectomy (P for nonlinear=9e-04) and total gastrectomy (P for nonlinear=0.029), but not for proximal gastrectomy (P for nonlinear=0.305). Nonlinear relation between progression free survival and tumor size can be seen from Figure 4. The largest increment of progression hazard was associated with tumor size larger than 5cm in the whole cohort(P for nonlinear=0.0407), proximal gastrectomy (P for nonlinear=0.0305),distal gastrectomy (P for nonlinear<0.001) and total gastrectomy (P for nonlinear<0.001).

**Figure 3.**
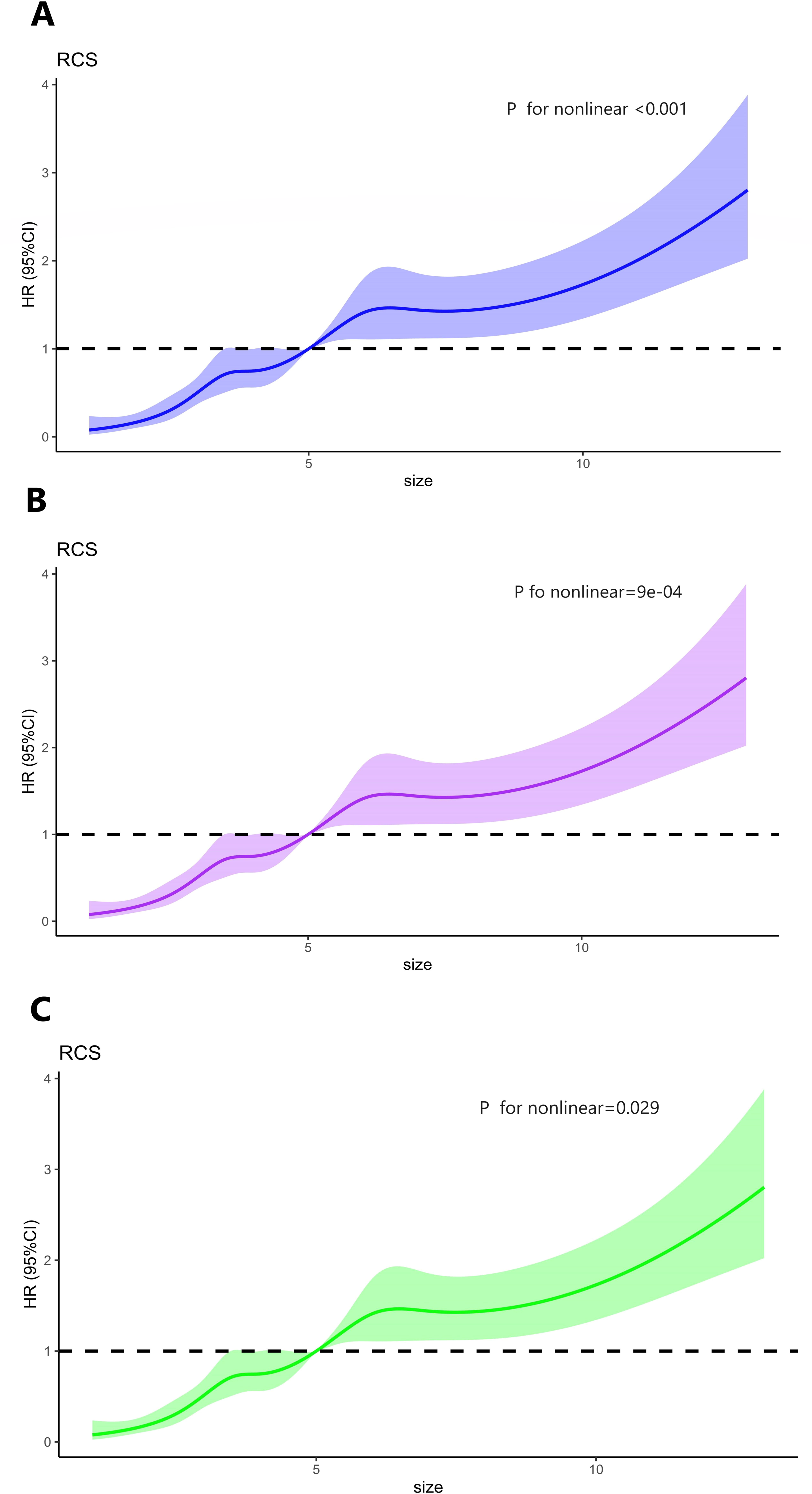
Restricted cubic splines for different tumor size of overall survival of gastric cancer patients underwent D2+gastrectomy. A: Whole cohort; B: Distal gastrectomy; C: Total gastrectomy

**Figure 4.**
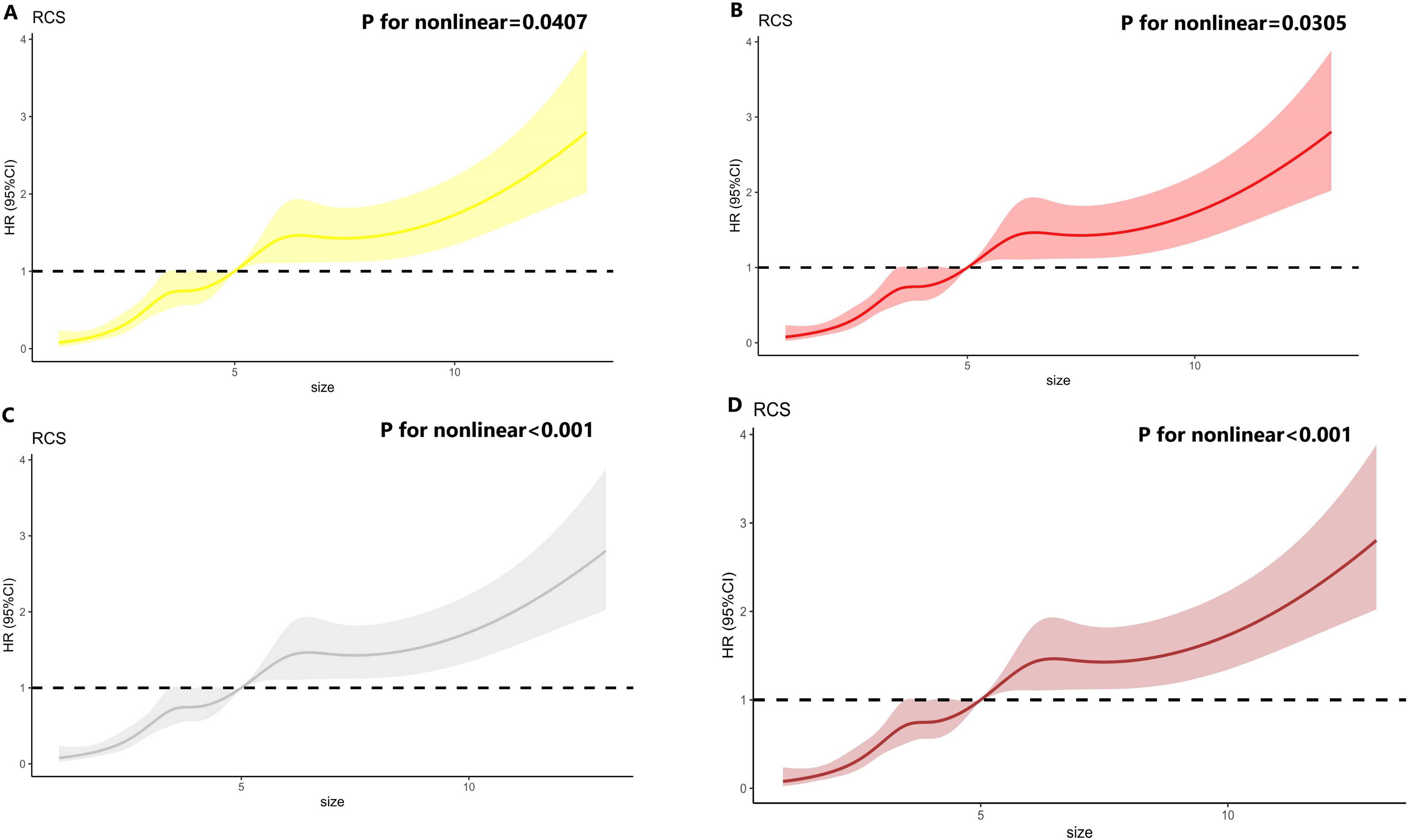
Restricted cubic splines for different tumor size of progression free survival of gastric cancer patients underwent D2+gastrectomy. A: Whole cohort; B: Proximal gastrectomy; C: Distal gastrectomy; D: Total gastrectomy

### Impact of Tumor Size with different tumor location on Survival

Different tumor location the relationships of tumor size and overall survival with different tumor site had been clearly presented in table 5. The outcomes demonstrated that the elevated mortality risk tightly related to tumor size for tumor located in upper 1/3(P for trend= 0.007) and the similar results(P for trend 0.045) still exhibited in further-adjusted model (adjusted^b^ model). However, the association between tumor size and mortality risk was not significant for tumor location of middle 1/3(P for trend= 0.452) and lower 1/3(P for trend= 0.102) in crude model, not to mention final adjusted model. The outcomes of progression free survival with different tumor size for upper 1/3, middle 1/3 and lower 1/3 seemed to be inconsistent in crude, adjusted^a^ and adjusted^b^ model(table 6). Tumor size was associated with progression risk in a size dependent manner (P trend = 0.008) for upper 1/3 in crude model, compared with the risk of those tumor size less than 2cm, the risk was significantly enhanced with tumor enlarged, whereas the result is not (P for nonlinear=0.056) in further-adjusted model (adjusted^b^ model). The similar finding showed in subgroup of lower 1/3 (crude model, P for trend= 0.023; adjusted^b^ model, P for trend= 0.123). In subgroup of middle 1/3, as more variables incorporated into model, the fully adjusted model achieved significative results(P for trend= 0.022).

In Figure 5 and Figure 6, restricted cubic splines(RCS) depicted that an irregularly shaped association between tumor size and the risk of death or progression. Nonlinear relationship identified in each cohorts, including upper 1/3, middle 1/3,lower 1/3 and whole cohort. When tumor larger than 5cm, the mortality and progression risk began to rise.

**Figure 5.**
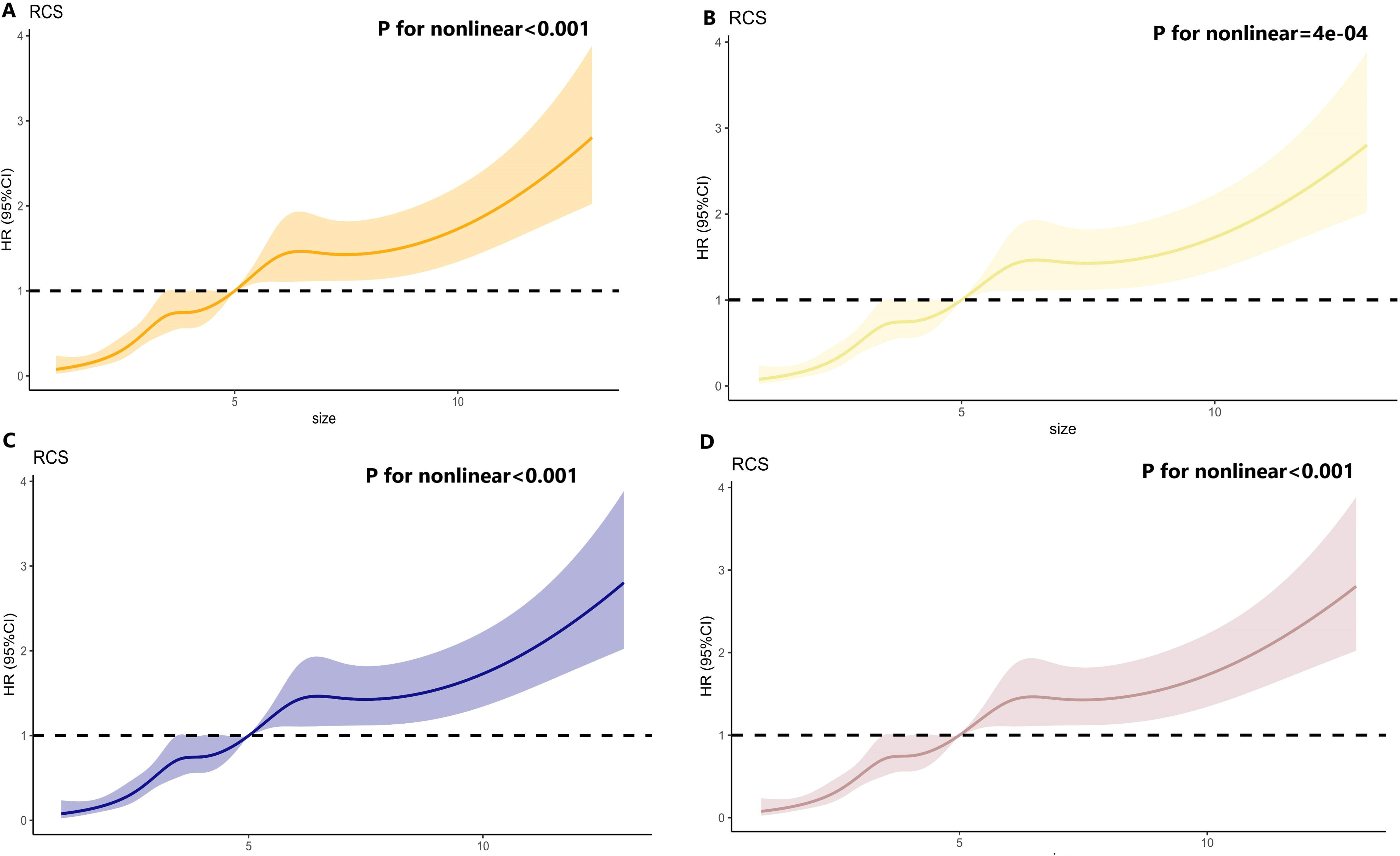
Restricted cubic splines for different tumor size of overall survival of gastric cancer patients with different tumor location. A: Whole cohort; B: Upper 1/3; C: Middle 1/3; D: Lower 1/3

**Figure 6.**
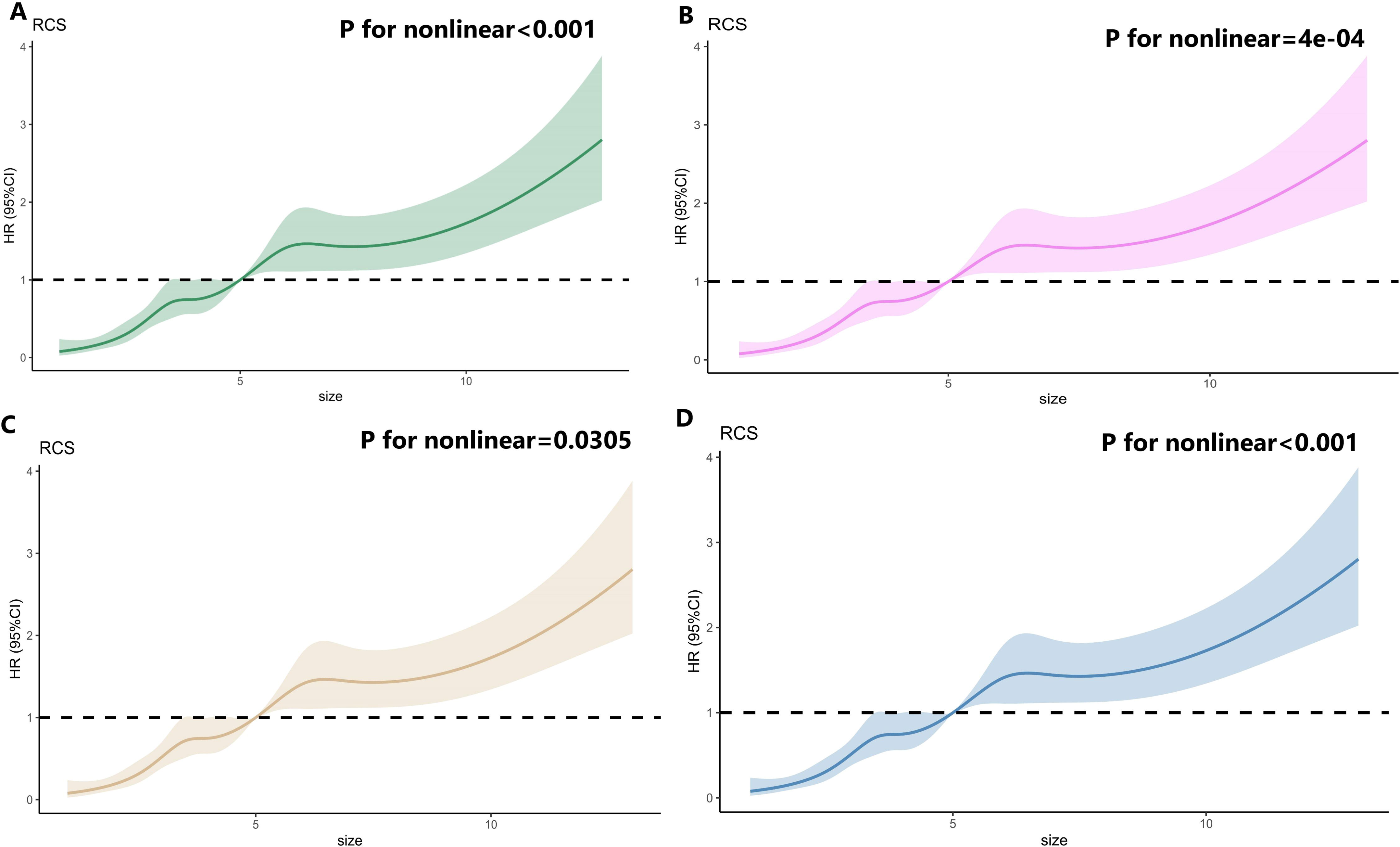
Restricted cubic splines for different tumor size of progression free survival of gastric cancer patients with different tumor location. A: Whole cohort; B: Upper 1/3; C: Middle 1/3; D: Lower 1/3

**Figure 7.**
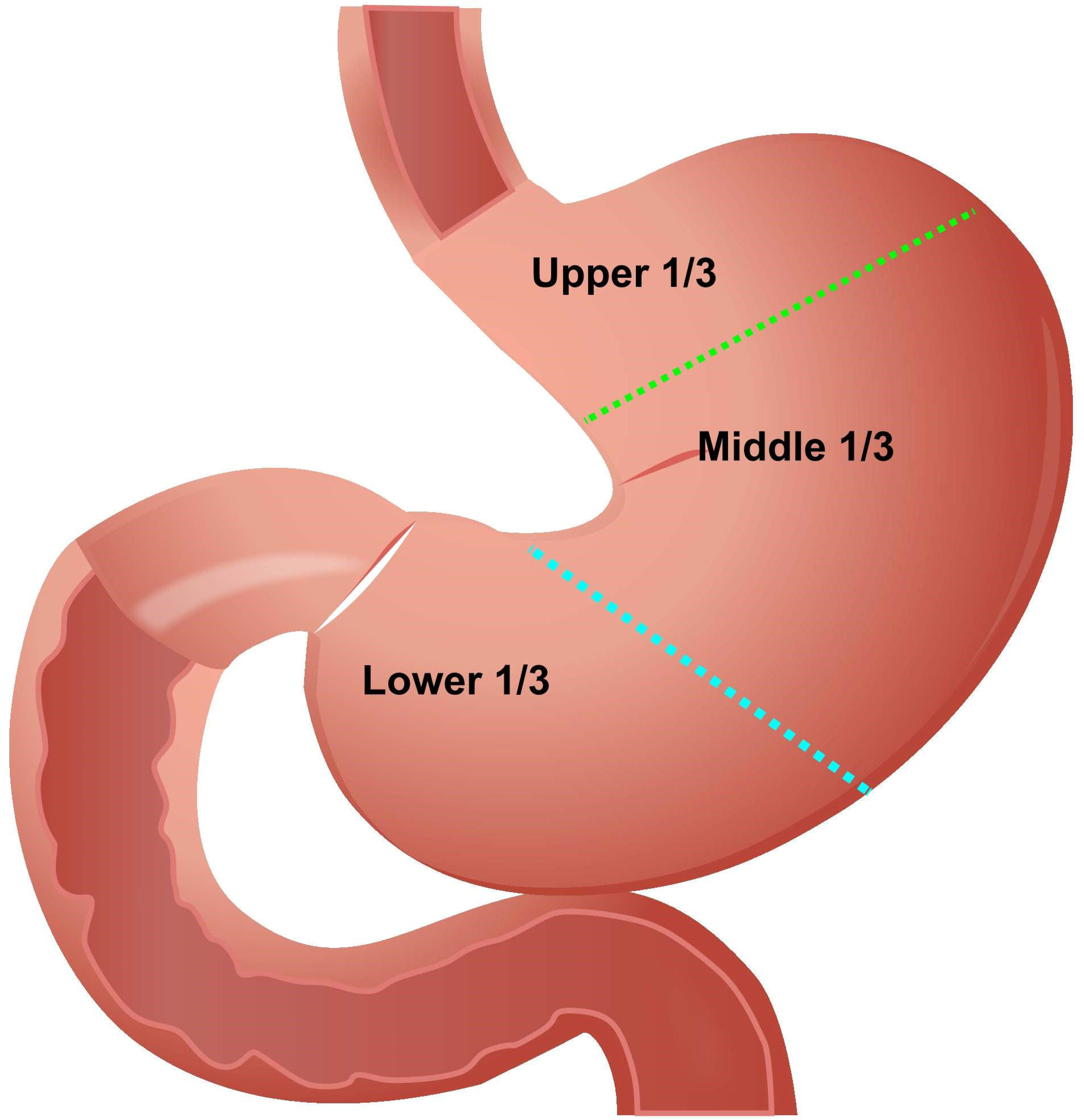
The division of different tumor location.

## Discussion

Gastric cancer is one of the main culprits affecting the incidence rate and mortality of cancer in China. And this study is currently the largest evaluative study conducted in China on the impact of tumor size and cancer survival rate ^[11]^.A relative study in Korean for patients who finished curative surgery for differentiated minute submucosal cancer, the final results demonstrated that one-dimensional tumor size > 2.9 cm and two-dimensional tumor size more than 8.3 cm2 have higher risk of postoperative morbidity and mortality^[12]^. Another study based on the SEER database showed that compared to the small size (≤ 2.5 cm) tumor group, the prognosis of the large size (≥ 5.3 cm) tumor group and the medium size (2.6-5.2 cm) tumor group was generally poor. However, there was no significant difference in OS survival between the moderate tumor group and the large tumor group ^[13]^. For patients with stage II or III gastric cancer, macroscopic tumor size (≥ 50 mm) is usually determined as an independent prognostic factor for patients undergoing S-1 adjuvant chemotherapy after surgery in China. However, Japanese institutional data mostly uses cancer patients with tumors<50 mm as research subjects. Compared to this, cancer research subjects in China have significantly higher rates of peritoneal or lymph node metastasis.^[14]^. Determining the size of tumors can help improve the accuracy of pT staging and prognosis prediction for cancer patients in China, especially for lymph node negative cancers^[15]^. According to research reports on Italian population data, patients with tumor size ≥ 2.5cm (p<0.01) tend to have better prognosis with tumor size<2.5cm, which is related to the depth of tumor invasion and lymph node metastasis ^[16]^. After D2 gastrectomy for T3-4aN0M0 cancer patients, it was found that the tumor size of the patient is directly related to the prognosis of adjuvant chemotherapy. However, in the patient population in China, patients with tumors larger than 5cm often exhibit poor survival rates.^[17]^.This analysis confirms that early diagnosis of cancer, pMMR, and high TNM staging of tumors are all beneficial for the recovery of cancer patients. Effective immunotherapy for dMMR and pMMR tumor subtypes has significant availability and can completely change the prospects of cancer treatment, including metastatic cancer. ^[18–20]^.

There are relatively few studies on the assessment of cancer recurrence based on tumor size, so at present, only lymph node metastasis can be used as the research basis. Through a study of 414 early-stage cancer patients, it was found that there was a general presence of small infiltration under the mucosa, posing a risk of stratified lymph node metastasis. The two-dimensional size of these patients’ tumors is>8.9 cm^2^, which also increases the risk of lymph node metastasis. ^[12]^. Another related study involved a total of 146 patients. They are all patients with early distal gastric cancer. This study aims to verify whether lymph node metastasis is related to tumor size, No.1, 3-7 lymph nodes metastasis more likely to occurtumor size <23 mm or≥23 mm and No.8a, 9 and 12a lymph nodes only arise in tumor size≥23 mm^[21]^. From the overall situation of the study, the impact of tumor size on the prognosis of patients is not very significant. However, there are significant differences in the different gastrectomy and tumor location. If the patients received proximal gastrectomy, it is positively correlated with the worst overall survival rate of the patient, and not directly related to progression free survival. However, for patients underwent distal or total gastrectomy, there is no significant difference between the tumor size and overall survival rate of patients, but the disparity of progression free survival between them still significant. Meanwhile, tumor located upper 1/3 with size >8cm related to worst overall survival rates compared to smaller tumor size, but the results of progression free survival is not consistent in crude model and fully adjusted model. Therefore, further research is still needed on how to reduce survival differences caused by tumor size.

After research, it has been confirmed that for patients who have completed D2+gastrectomy, especially those who undergo chemotherapy before or after surgery, tumor size can serve as a good prognostic factor. When doctors determine the adjuvant treatment plan for patients with different tumor location and experienced D2+gastrectomy, although most do not take tumor size into account, the use of four types of tumor size presents an equal distribution: 2-4cm, 4-6cm, 6-8cm and >8cm, except for <2cm. Tumor size>5cm was regard as a line of demarcation of mortality and progression, that results from restricted cubic splines(RCS) showed. When tumor size larger than 5 cm, mortality and progression hazard began to ascend, except for patients received proximal gastrectomy, that means postoperative treatment should be focused on for those patients. It is worth noting that for patients receiving adjuvant chemotherapy after proximal gastrectomy and tumor located upper 1/3, their classification of tumor size will significantly show overall survival differences, the similar effect occurred in progression free survival of patients accepted distal, total gastrectomy. The results of this study showed that tumors larger than 8cm in patients directly affect the overall survival of proximal gastrectomy, tumor located upper 1/3 and progression free survival of distal and total gastrectomy, while tumor great than 5 cm showed a significant impact in each group, but not for proximal gastrectomy.

The reason of complicated prognostic effect of different tumor location associated with progression free survival may be elucidated as following. On one hand, the scope of tumor size located middle 1/3 was so narrow that the large size groups(6-8cm and >8cm) was slip out of this stage. On the other hand, the final results of overall survival was death and progression free survival was death or recurrence. In view of this, the disparity of overall survival and progression free survival with different tumor location owing to the difference of recurrence between those groups. More palindromic patients enrolled in the final results of progression free survival was principally responsible for the reversed results of tumor location of OS and PFS.

There were several limitations in the current study. Firstly, the members participating in the statistical analysis are all from the same medical center and need to undergo relevant validation from other medical centers; Second, larger sample size of different tumor location associated with progression free survival is needed to confirm or exclude prognostic effects of recurrence and metastasis.

Despite the limitations mentioned above, this study is the first to evaluate the impact of tumor size on cancer prognosis and predictive value, especially for domestic cancer patients after different gastrectomy and the associations between different tumor locations. This study found that for patients after D2+ gastrectomy, tumor size directly affects the prognostic value in different way for different group. Moreover, we can conclude that for gastric cancer patients whose tumor greater than 5cm associated with more chance of mortality and progression except for subgroup of proximal gastrectomy, and if the tumor is larger than 8cm, the prognosis of the patients is generally poor.

## Conclusions

Tumor size>5cm can be seen as a line of demarcation of mortality and progression of gastric cancer after D2+gastrectomy, the hazard ratio began to rise when tumor size large than 5cm. Further research is needed by relevant scholars on the correlation between tumor size and patient prognosis staging models, as well as how to incorporate research results into relevant cancer treatment decisions.

## Acknowledgments

Special thanks were given to Suwen Ji, Chenjie Fan for proofreading the patient follow-up information.

## Ethics approval

This research study was conducted retrospectively from data obtained for clinical purposes. We consulted extensively with Ethics Committee of Shanxi Cancer Hospital and granted ethical approval from the ethics Committee of Shanxi Carcinoma Hospital (No:2022JC23). All methods were performed in accordance with the relevant guidelines and regulations. All procedures performed in studies involving human participants were in accordance with the ethical standards of our institutional research committee.

## Consent to participate

Not applicable.

## Consent for publication

Not applicable.

## Data availability

The raw datasets generated during the current study are available from the corresponding author on reasonable request. All data analyzed during this study are included in this published article, Yifan Li is willing to share my data in this article.

## Authors’ Contributions

Yifan Li wrote the main manuscript text, prepared figures 1–7, Haoliang Zhao, Yinan Shi, Ruirui Yang reviewed and revised the manuscript.

