## Supplementary material for "Tumor size>5cm is a line of demarcation of mortality and progression of gastric cancer after D2+gastrectomy for Chinese population": Table

Table1. Demographics of study population

| Variables | Total (n=1708) | stageⅠ(n=336) | stageⅡ(n=412) | stage Ⅲ(n=902) | stageⅣ(n=58) |
| --- | --- | --- | --- | --- | --- |
| Gender |  |  |  |  |  |
| Male | 1358(79.5%) | 271(80.7%) | 340(82.5%) | 704(78.0%) | 43(74.1%) |
| Female | 350(20.5%) | 65(19.3%) | 72(17.5%) | 198(22.0%) | 15(25.9%) |
| Age (years) |  |  |  |  |  |
| 20-49 | 336(19.7%) | 63(18.8%) | 77(18.7%) | 180(20.0%) | 16(27.6%) |
| 50-59 | 566(33.1%) | 124(36.9%) | 128(36.9%) | 294(32.6%) | 20(34.5%) |
| 60-69 | 624(36.5%) | 117(34.8%) | 167(40.5%) | 323(35.8%) | 17(29.3%) |
| ≥70 | 182(10.7%) | 32(9.5%) | 40(9.7%) | 105(11.6%) | 5(8.6%) |
| pT stage |  |  |  |  |  |
| T1 | 306(17.9%) | 291(86.6%) | 13(3.2%)) | 0 | 2(3.4%) |
| T2 | 76(4.4%) | 45(13.4%) | 29(7.0%) | 2(0.2%) | 1(1.7%) |
| T3 | 503(29.4%) | 0 | 274(66.5%) | 224(24.8%) | 4(6.9%) |
| T4 | 823(48.2%) | 0 | 96(23.3%) | 676(74.9%) | 51(87.9%) |
| Number of positive lymph nodes | |  |  |  |  |
| 0 | 563(33.0%) | 317(94.3%) | 216(38.4%) | 110(19.5%) | 44(7.8%) |
| 1-2 | 336(19.7%) | 19(5.7%) | 155(46.1%) | 110(32.7%) | 34(10.1%) |
| 3-6 | 240(14.1%) | 0 | 74(30.8%) | 93(38.8%) | 49(20.4%) |
| ≥7 | 569(33.3%) | 0 | 117(20.6%) | 215(37.8%) | 135(23.7%) |
| Tumor location |  |  |  |  |  |
| Upper | 866(50.7%) | 140(41.7%) | 219(53.2%) | 478(53.0%) | 29(50.0%) |
| Middle | 282(16.5%) | 43(12.8%) | 64(15.5%) | 163(18.1%) | 12(20.7%) |
| Lower | 547(32.0%) | 148(44.0%) | 127(30.8%) | 255(28.3%) | 17(29.3%) |
| Multiple | 13(0.8%) | 5(1.5%) | 2(0.5%) | 6(0.7%) | 0 |
| Vascular invasion |  |  |  |  |  |
| Negative | 784(45.9%) | 307(91.4%) | 245(59.5%) | 221(24.5%) | 11(19.0%) |
| Positive | 924(54.1%) | 29(8.6%) | 167(40.5%) | 681(75.5%) | 47(81.0%) |
| Neural invasion |  |  |  |  |  |
| Negative | 904(52.9%) | 325(96.7%) | 255(61.9%) | 306(33.9%) | 18(31.0%) |
| Positive | 804(47.1%) | 11(3.3%) | 155(38.1%) | 596(66.1%) | 40(69.0%) |
| Lauren classification | |  |  |  |  |
| Intestinal | 670(39.2%) | 274(81.5%) | 203(49.3%) | 177(19.6%) | 16(27.6%) |
| Diffuse | 608(35.6%) | 24(7.1%) | 85(20.6%) | 468(51.9%) | 31(53.4%) |
| Mixed | 430(25.2%) | 38(11.3%) | 124(30.1%) | 257(28.5%) | 11(19.0%) |
| Type of gastrectomy |  |  |  |  |  |
| Proximal | 166(9.7%) | 73(21.7%) | 38(9.2%) | 52(5.8%) | 3(5.2%) |
| Distal | 554(32.4%) | 164(48.8%) | 134(32.5%) | 243(26.9%) | 13(22.4%) |
| Total | 986(57.7%) | 97(28.9%) | 240(58.3%) | 607(67.3%) | 42(72.4%) |
| PPG | 2(0.1%) | 2(0.6%) | 0 | 0 | 0 |
| Omentum metastasis |  |  |  |  |  |
| Negative | 1661(97.2%) | 335(99.7%) | 412(100%) | 860(95.3%) | 54(93.1%) |
| Positive | 47(2.8%) | 1(0.3%) | 0 | 42(4.7%) | 4(6.9%) |
| Surgical margin |  |  |  |  |  |
| Negative | 1626(95.2%) | 329(97.9%) | 404(98.1%) | 837(92.8%) | 56(96.6%) |
| Positive | 82(4.8%) | 7(2.1%) | 8(1.9%) | 65(7.2%) | 2(3.4%) |
| Histological classification | |  |  |  |  |
| Adenocarcinoma | 1413(82.7%) | 301(89.6%) | 344(83.5%) | 721(79.9%) | 47(81.0%) |
| Others | 295(17.3%) | 35(10.4%) | 68(16.5%) | 181(20.1%) | 11(19.0%) |
| Her-2 |  |  |  |  |  |
| Negative | 1051(61.5%) | 236(70.2%) | 241(58.5%) | 533(59.1%) | 158(15.0%) |
| Positive | 657(38.5%) | 100(29.8%) | 171(41.5%) | 369(40.9%) | 104(15.8%) |
| AE1/AE3 |  |  |  |  |  |
| Negative | 370(21.7%) | 231(68.8%) | 86(20.9%) | 51(5.7%) | 2(3.4%) |
| Positive | 1338(78.3%) | 105(31.2%) | 326(79.1%) | 851(94.3%) | 56(96.6%) |
| CK7 |  |  |  |  |  |
| Negative | 868(50.8%) | 165(49.1%) | 212(51.5%) | 460(51.0%) | 31(53.4%) |
| Positive | 840(49.2%) | 171(50.9%) | 200(48.5%) | 442(49.0%) | 27(46.6%) |
| CK20 |  |  |  |  |  |
| Negative | 1239(72.5%) | 220(65.5%) | 314(76.2%) | 662(73.4%) | 43(74.1%) |
| Positive | 469(27.5%) | 116(34.5%) | 98(23.8%) | 240(26.6%) | 15(25.9%) |
| CDX-2 |  |  |  |  |  |
| Negative | 960(56.2%) | 249(74.1%) | 220(53.4%) | 459(50.9%) | 32(55.2%) |
| Positive | 748(43.8%） | 87(25.9%) | 192(46.6%) | 443(49.1%) | 26(44.8%) |
| SATB-2 |  |  |  |  |  |
| Negative | 1400(82.0%) | 295(87.8%) | 349(84.7%) | 711(78.8%) | 45(77.6%) |
| Positive | 308(18.0%) | 41(12.2%) | 63(15.3%) | 191(21.2%) | 13(22.4%) |
| SYN |  |  |  |  |  |
| Negative | 1265(73.5%) | 261(77.7%) | 311(75.5%) | 643(71.3%) | 41(70.7%) |
| Positive | 452(26.5%) | 75(22.3%) | 101(24.5%) | 259(28.7%) | 17(29.3%) |
| CGA |  |  |  |  |  |
| Negative | 1397(81.8%) | 240(71.4%) | 344(83.5%) | 767(85.0%) | 46(79.3%) |
| Positive | 311(18.2%) | 96(38.6%) | 68(16.5%) | 135(15.0%) | 12(20.7%) |
| CD56 |  |  |  |  |  |
| Negative | 1090(63.8%) | 291(86.6%) | 237(57.5%) | 530(58.8%) | 32(55.2%) |
| Positive | 618(36.2%) | 45(13.4%) | 175(42.5%) | 372(41.2%) | 26(44.8%) |
| MLH1 |  |  |  |  |  |
| Negative | 218(12.8%) | 121(36.0%) | 46(11.2%) | 49(5.4%) | 2(3.4%) |
| Positive | 1490(87.2%) | 215(64.0%) | 366(88.8%) | 853(94.6%) | 56(96.6%) |
| PMS2 |  |  |  |  |  |
| Negative | 467(27.3%) | 198(58.9%) | 102(24.8%) | 161(17.8%) | 6(10.3%) |
| Positive | 1241(72.7%) | 138(41.1%) | 310(75.2%) | 741(82.2%) | 52(89.7%) |
| MSH2 |  |  |  |  |  |
| Negative | 232(13.6%) | 133(39.6%) | 39(9.5%) | 55(6.1%) | 5(8.6%) |
| Positive | 1476(86.4%) | 203(60.4%) | 373(90.5%) | 847(93.9%) | 53(91.4%) |
| MSH6 |  |  |  |  |  |
| Negative | 219(12.8%) | 136(40.5%) | 33(8.0%) | 46(5.1%) | 4(6.9%) |
| Positive | 1489(87.2%) | 200(59.5%) | 379(92.0%) | 856(94.9%) | 54(93.1%) |

Table2. Univariate correlation with tumor size

| Variables | Total (n=1708) | 0-2cm (n=206) | 2-4cm (n=562) | 4-6cm (n=528) | 6-8cm (n=262) | >8cm (n=150) | P |
| --- | --- | --- | --- | --- | --- | --- | --- |
| Gender |  |  |  |  |  |  | 0.356 |
| Male | 1358 | 167(12.3%) | 434(32.0%) | 433(31.9%) | 205(15.1%) | 119(8.8%) |  |
| Female | 350 | 39(11.1%) | 128(36.6%) | 95(27.1%) | 57(16.3%) | 31(8.9%) |  |
| Age (years) |  |  |  |  |  |  | 0.257 |
| 20-49 | 336 | 36(10.7%) | 127(37.8%) | 94(28.0%) | 49(14.6%) | 30(8.9%) |  |
| 50-59 | 566 | 80(14.1%) | 192(33.9%) | 167(29.5%) | 85(15.0%) | 42(7.4%) |  |
| 60-69 | 624 | 70(11.2%) | 186(29.8%) | 212(34.0%) | 100(16.0%) | 56(9.0%) |  |
| ≥70 | 182 | 20(11.0%) | 57(31.3%) | 55(30.2%) | 28(15.4%) | 28(15.4%) |  |
| pT stage |  |  |  |  |  |  | <0.001 |
| T1 | 306 | 163(53.3%) | 115(37.6%) | 22(7.2%) | 3(1.0%) | 3(1.0%) |  |
| T2 | 76 | 8(10.5%) | 46(60.5%) | 15(19.7%) | 5(6.6%) | 2(2.6%) |  |
| T3 | 503 | 22(4.4%) | 188(37.4%) | 184(36.6%) | 73(14.5%) | 36(7.2%) |  |
| T4 | 823 | 13(1.6%) | 213(25.9%) | 307(37.3%) | 181(22.0%) | 109(13.2%) |  |
| Number of positive lymph nodes | |  |  |  |  |  | <0.001 |
| 0 | 563 | 176(31.3%) | 216(38.4%) | 110(19.5%) | 44(7.8%) | 17(3.0%) |  |
| 1-2 | 336 | 16(4.8%) | 155(46.1%) | 110(32.7%) | 34(10.1%) | 21(6.3%) |  |
| 3-6 | 240 | 8(3.3%) | 74(30.8%) | 93(38.8%) | 49(20.4%) | 16(6.7%) |  |
| ≥7 | 569 | 6(1.1%) | 117(20.6%) | 215(37.8%) | 135(23.7%) | 96(16.9%) |  |
| pTNM stage |  |  |  |  |  |  | <0.001 |
| Ⅰ | 336 | 168(50.0%) | 135(40.2%) | 25(7.4%) | 5(1.5%) | 3(0.9%) |  |
| Ⅱ | 412 | 23(5.6%) | 173(42.0%) | 143(34.7%) | 53(12.9%) | 20(4.9%) |  |
| Ⅲ | 902 | 14(1.6%) | 240(26.6%) | 337(37.4%) | 193(21.4%) | 118(13.1%) |  |
| Ⅳ | 58 | 1(1.7%) | 14(24.1%) | 23(39.7%) | 11(19.0%) | 9(15.5%) |  |
| Tumor location |  |  |  |  |  |  | 0.002 |
| Upper | 866 | 86(9.9%) | 252(29.1%) | 296(34.2%) | 153(17.7%) | 79(9.1%) |  |
| Middle | 282 | 32(11.3%) | 80(28.4%) | 66(23.4%) | 57(20.2%) | 47(16.7%) |  |
| Lower | 547 | 88(16.1%) | 224(41.0%) | 161(29.4%) | 50(9.1%) | 24(4.4%) |  |
| Multiple | 13 | 0 | 6(46.2%) | 5(38.5%) | 2(15.4%) | 0 |  |
| Vascular  invasion |  |  |  |  |  |  | <0.001 |
| Negative | 784 | 181(23.1%) | 289(36.9%) | 191(24.4%) | 72(9.2%) | 51(6.5%) |  |
| Positive | 924 | 25(2.7%) | 273(29.5%) | 337(36.5%) | 190(20.6%) | 99(10.7%) |  |
| Neural invasion |  |  |  |  |  |  | <0.001 |
| Negative | 904 | 180(19.9%) | 336(37.2%) | 232(25.7%) | 100(11.1%) | 56(6.2%) |  |
| Positive | 804 | 26(3.2%) | 226(28.1%) | 296(36.8%) | 162(20.1%) | 94(11.7%) |  |
| Lauren  classification |  |  |  |  |  |  | <0.001 |
| Intestinal | 670 | 155(23.1%) | 245(36.5%) | 171(25.5%) | 70(10.4%) | 29(4.3%) |  |
| Diffuse | 608 | 24(3.9%) | 158(26.0%) | 207(34.0%) | 134(22.0%) | 85(14.0%) |  |
| Mixed | 430 | 27(6.3%) | 159(37.0%) | 150(34.9%) | 58(13.5%) | 36(8.4%) |  |
| Type of gastrectomy | |  |  |  |  |  | <0.001 |
| Proximal | 166 | 46(27.7%) | 68(41.0%) | 40(24.1%) | 7(4.2%) | 5(3.0%) |  |
| Distal | 554 | 104(18.8%) | 230(41.5%) | 162(29.2%) | 42(7.6%) | 16(2.9%) |  |
| Total | 986 | 54(5.5%) | 264(26.8%) | 326(33.1%) | 213(21.6%) | 129(13.1%) |  |
| PPG | 2 | 2(100%) | 0 | 0 | 0 | 0 |  |
| Omentum metastasis | |  |  |  |  |  | <0.001 |
| Negative | 1661 | 206(12.4%) | 554(33.4%) | 514(30.9%) | 251(15.1%) | 136(8.2%) |  |
| Positive | 47 | 0 | 8(17.0%) | 14(29.8%) | 11(23.4%) | 14(29.8%) |  |
| Surgical margin |  |  |  |  |  |  | <0.001 |
| Negative | 1626 | 201(12.4%) | 549(33.8%) | 500(30.8%) | 244(15.0%) | 132(8.1%) |  |
| Positive | 82 | 5(6.1%) | 13(15.9%) | 28(24.1%) | 18(22.0%) | 18(22.0%) |  |
| Histological classification | |  |  |  |  |  | 0.001 |
| Adenocarcinoma | 1413 | 188(13.3%) | 473(33.5%) | 430(30.4%) | 206(14.6%) | 116(8.2%) |  |
| Others | 295 | 18(6.1%) | 89(30.2%) | 98(33.2%) | 56(19.0%) | 34(11.5%) |  |
| Her-2 |  |  |  |  |  |  | 0.058 |
| Negative | 1051 | 143(13.6%) | 353(33.7%) | 311(29.6%) | 158(15.0%) | 85(8.1%) |  |
| Positive | 657 | 63(9.6%) | 208(31.7%) | 217(33.0%) | 104(15.8%) | 65(9.9%) |  |
| AE1/AE3 |  |  |  |  |  |  | <0.001 |
| Negative | 370 | 137(37.0%) | 139(37.6%) | 73(19.7%) | 14(3.8%) | 7(1.9%) |  |
| Positive | 1338 | 69(5.2%) | 423(31.6%) | 455(34.0%) | 248(18.5%) | 143(10.7%) |  |
| CK7 |  |  |  |  |  |  | 0.632 |
| Negative | 868 | 101(11.6%) | 273(31.5%) | 279(32.1%) | 137(15.8%) | 78(9.0%) |  |
| Positive | 840 | 105(51.0%) | 289(51.4%) | 249(47.2%) | 125(47.7%) | 72(48.0%) |  |
| CK20 |  |  |  |  |  |  | 0.062 |
| Negative | 1239 | 132(10.7%) | 417(33.7%) | 386(31.2%) | 196(15.8%) | 108(8.7%) |  |
| Positive | 469 | 74(15.8%) | 145(30.9%) | 142(30.3%) | 66(14.1%) | 42(9.0%) |  |
| CDX-2 |  |  |  |  |  |  | <0.001 |
| Negative | 960 | 156(16.3%) | 308(32.1%) | 291(30.3%) | 124(12.9%) | 81(8.4%) |  |
| Positive | 748 | 50(6.7%) | 254(34.0%) | 237(31.7%) | 138(18.4%) | 69(9.2%) |  |
| SATB-2 |  |  |  |  |  |  | 0.066 |
| Negative | 1400 | 181(12.9%) | 468(33.4%) | 421(30.1%) | 207(14.8%) | 123(8.8%) |  |
| Positive | 308 | 25(8.1%) | 94(30.5%) | 107(34.7%) | 55(17.9%) | 27(8.8%) |  |
| SYN |  |  |  |  |  |  | 0.016 |
| Negative | 1265 | 164(10.9%) | 426(33.9%) | 386(30.7%) | 180(14.3%) | 100(8.0%) |  |
| Positive | 452 | 42(9.3%) | 136(30.1%) | 142(31.4%) | 82(18.1%) | 50(11.1%) |  |
| CGA |  |  |  |  |  |  | 0.006 |
| Negative | 1397 | 152(10.9%) | 455(32.6%) | 447(32.0%) | 223(16.0%) | 120(8.6%) |  |
| Positive | 311 | 54(7.4%) | 107(34.4%) | 81(26.0%) | 39(12.5%) | 30(9.6%) |  |
| CD56 |  |  |  |  |  |  | <0.001 |
| Negative | 1090 | 177(16.2%) | 359(32.9%) | 315(28.9%) | 155(14.2%) | 84(7.7%) |  |
| Positive | 618 | 29(4.7%) | 203(32.8%) | 213(34.5%) | 107(17.3%) | 66(10.7%) |  |
| MLH1 |  |  |  |  |  |  | <0.001 |
| Negative | 218 | 81(37.2%) | 72(33.0%) | 37(17.0%) | 16(7.3%) | 12(5.5%) |  |
| Positive | 1490 | 125(8.4%) | 490(32.9%) | 491(33.0%) | 246(16.5%) | 138(9.3%) |  |
| PMS2 |  |  |  |  |  |  | <0.001 |
| Negative | 467 | 121(25.9%) | 161(34.5%) | 110(23.6%) | 47(10.1%) | 28(6.0%) |  |
| Positive | 1241 | 85(6.8%) | 401(32.3%) | 418(33.7%) | 215(17.3%) | 122(9.8%) |  |
| MSH2 |  |  |  |  |  |  | <0.001 |
| Negative | 232 | 77(33.2%) | 86(37.1%) | 41(17.7%) | 16(6.9%) | 12(5.2%) |  |
| Positive | 1476 | 129(8.7%) | 476(32.2%) | 487(33.0%) | 246(16.7%) | 138(9.3%) |  |
| MSH6 |  |  |  |  |  |  | <0.001 |
| Negative | 219 | 88(40.2%) | 77(35.2%) | 29(13.2%) | 17(7.8%) | 8(3.7%) |  |
| Positive | 1489 | 118(7.9%) | 485(32.6%) | 499(33.5%) | 245(16.5%) | 142(9.5%) |  |

Table 3 Crude and adjusted hazard ratios estimates for overall survival(OS) with different tumor size by each types of gastrectomy

| Types of gastrectomy | 0-2cm | 2-4cm | 4-6cm | 6-8cm | >8cm | *P* for trend |
| --- | --- | --- | --- | --- | --- | --- |
| Proximal |  |  |  |  |  |  |
| Crude HR(95%CI) | 1.000 | 3.928(0.789-19.547) | 3.605(0.646-20.106) | 15.092(2.230-102.136) | 15.253(1.984-117.263) | 0.013 |
| Adjusted^a^ HR(95%CI) | 1.000 | 3.801(0.762-18.963) | 3.750(0.652-21.564) | 15.513(2.254-106.78) | 14.945(1.871-119.375) | 0.017 |
| Adjusted^b^ HR(95%CI) | 1.000 | 5.504(0.871-29.313) | 4.38(0.636-30.193) | 41.205(4.339-391.334) | 36.253(3.411-385.344) | 0.002 |
| Distal |  |  |  |  |  |  |
| Crude HR(95%CI) | 1.000 | 2.126(0.869-5.201) | 2.731(1.106-6.739) | 2.163(0.789-5.929) | 3.683(1.241-10.937) | 0.112 |
| Adjusted^a^ HR(95%CI) | 1.000 | 2.237(0.909-5.508) | 2.827(1.141-7.004) | 2.255(0.816-6.234) | 3.795(1.269-11.352) | 0.120 |
| Adjusted^b^ HR(95%CI) | 1.000 | 2.087(0.837-5.204) | 2.812(1.113-7.109) | 2.108(0.754-5.896) | 3.506(1.151-10.678) | 0.120 |
| Total |  |  |  |  |  |  |
| Crude HR(95%CI) | 1.000 | 1.436(0.544-3.792) | 1.812(0.687-4.782) | 1.928(0.725-5.123) | 2.172(0.812-5.814) | 0.102 |
| Adjusted^a^ HR(95%CI) | 1.000 | 1.435(0.546-3.772) | 1.84(0.7-4.834) | 1.853(0.7-4.903) | 2.164(0.811-5.775) | 0.126 |
| Adjusted^b^ HR(95%CI) | 1.000 | 1.505(0.57-3.974) | 1.898(0.718-5.018) | 2.003(0.753-5.329) | 2.25(0.836-6.054) | 0.132 |

Crude HR is adjusted for age at recruitment, sex, pT stage, Number of positive lymph nodes.

Adjusted^a^ HR is adjusted for the variables in Crude HR model plus nerve invasion, vascular invasion, surgical margin, omentum metastasis, Lauren classification

Adjusted^b^ HR is adjusted for all variables in Adjusted^a^ model , plus AE1/AE3, CD56, CDX-2, CGA, CK7, CK20, SATB-2, SYN, PMS2, MLH1, MSH2 MSH6

Table 4 Crude and adjusted hazard ratios estimates for progression free survival (PFS) with different tumor size by each type of gastrectomy

| Types of gastrectomy | 0-2cm | 2-4cm | 4-6cm | 6-8cm | >8cm | *P* for trend |
| --- | --- | --- | --- | --- | --- | --- |
| Proximal |  |  |  |  |  |  |
| Crude HR(95%CI) | 1.000 | 4.97(0.994-24.852) | 4.105(0.749-22.51) | 7.085(1.039-48.31) | 7.207(0.965-53.81) | 0.259 |
| Adjusted^a^ HR(95%CI) | 1.000 | 4.744(0.944-23.85) | 3.413(0.603-19.33) | 6.837(0.974-48.01) | 6.595(0.856-50.84) | 0.237 |
| Adjusted^b^ HR(95%CI) | 1.000 | 5.497(0.941-31.11) | 2.835(0.454-17.70) | 24.55(2.58-234.01) | 9.007(0.986-82.296) | 0.021 |
| Distal |  |  |  |  |  |  |
| Crude HR(95%CI) | 1.000 | 1.114(0.594-2.088) | 1.324(0.693-2.53) | 1.162(0.548-2.467) | 3.093(1.332-7.182) | 0.029 |
| Adjusted^a^ HR(95%CI) | 1.000 | 1.143(0.604-2.161) | 1.32(0.684-2.546) | 1.297(0.606-2.779) | 3.212(1.364-7.561) | 0.037 |
| Adjusted^b^ HR(95%CI) | 1.000 | 1.101(0.572-2.12) | 1.256(0.638-2.473) | 1.335(0.615-2.901) | 3.322(1.322-8.501) | 0.03 |
| Total |  |  |  |  |  |  |
| Crude HR(95%CI) | 1.000 | 1.516(0.626-1.044) | 1.928(0.798-4.661) | 2.159(0.887-5.253) | 2.506(1.023-6.139) | 0.011 |
| Adjusted^a^ HR(95%CI) | 1.000 | 1.553(0.643-3.753) | 1.966(0.813-4.755) | 2.162(0.888-5.263) | 2.586(1.05-6.373) | 0.018 |
| Adjusted^b^ HR(95%CI) | 1.000 | 0.736(0.369-1.467) | 0.892(0.408-1.949) | 0.425(0.091-1.997) | 2.174(0.725-6.516) | 0.016 |

Crude HR is adjusted for age at recruitment, sex, pT stage, Number of positive lymph nodes.

Adjusted^a^ HR is adjusted for the variables in Crude HR model plus nerve invasion, vascular invasion, surgical margin, omentum metastasis, Lauren classification

Adjusted^b^ HR is adjusted for all variables in Adjusted^a^ model , plus AE1/AE3, CD56, CDX-2, CGA, CK7, CK20, SATB-2, SYN, PMS2, MLH1, MSH2 MSH6

Table 5 Crude and adjusted hazard ratios estimates for overall survival(OS) with different tumor size by each tumor location

| Tumor location | 0-2cm | 2-4cm | 4-6cm | 6-8cm | >8cm | *P* for trend |
| --- | --- | --- | --- | --- | --- | --- |
| Upper 1/3 |  |  |  |  |  |  |
| Crude HR(95%CI) | 1.000 | 3.869(1.102-13.588) | 5.186(1.459-18.442) | 6.181(1.722-22.178) | 6.202(1.699-22.638) | 0.007 |
| Adjusted^a^ HR(95%CI) | 1.000 | 3.916(1.188-13.722) | 5.166(1.456-18.333) | 5.831(1.626-20.906) | 5.938(1.628-21.633) | 0.02 |
| Adjusted^b^ HR(95%CI) | 1.000 | 3.68(1.037-13.065) | 4.695(1.301-16.943) | 5.294(1.454-19.266) | 5.338(1.439-19.805) | 0.045 |
| Middle 1/3 |  |  |  |  |  |  |
| Crude HR(95%CI) | 1.000 | 1.708(0.447-6.523) | 2.731(0.419-6.441) | 1.582(0.401-6.24) | 2.331(0.597-9.10) | 0.452 |
| Adjusted^a^ HR(95%CI) | 1.000 | 1.728(0.448-6.662) | 1.797(0.455-7.103) | 1.708(0.431-6.768) | 2.821(0.715-11.134) | 0.231 |
| Adjusted^b^ HR(95%CI) | 1.000 | 2.052(0.506-8.323) | 2.538(0.611-10.544) | 2.019(0.482-8.468) | 3.884(0.932-16.179) | 0.089 |
| Lower 1/3 |  |  |  |  |  |  |
| Crude HR(95%CI) | 1.000 | 1.677(0.726-3.873) | 2.152(0.924-5.008) | 2.05(0.814-5.166) | 2.716(1.035-7.129) | 0.102 |
| Adjusted^a^ HR(95%CI) | 1.000 | 1.623(0.694-3.797) | 2.114(0.897-4.981) | 2.06(0.807-5.262) | 2.358(0.869-6.399) | 0.126 |
| Adjusted^b^ HR(95%CI) | 1.000 | 1.583(0.666-3.763) | 2.222(0.921-5.357) | 2.054(0.791-5.335) | 2.034(0.731-5.664) | 0.285 |

Crude HR is adjusted for age at recruitment, sex, pT stage, Number of positive lymph nodes.

Adjusted^a^ HR is adjusted for the variables in Crude HR model plus nerve invasion, vascular invasion, surgical margin, omentum metastasis, Lauren classification

Adjusted^b^ HR is adjusted for all variables in Adjusted^a^ model , plus AE1/AE3, CD56, CDX-2, CGA, CK7, CK20, SATB-2, SYN, PMS2, MLH1, MSH2 MSH6

Table 6 Crude and adjusted hazard ratios estimates for progression free survival (PFS) with different tumor size by each tumor location

| Tumor location | 0-2cm | 2-4cm | 4-6cm | 6-8cm | >8cm | *P* for trend |
| --- | --- | --- | --- | --- | --- | --- |
| Upper 1/3 |  |  |  |  |  |  |
| Crude HR(95%CI) | 1.000 | 3.209(1.064-9.679) | 4.091(1.343-12.46) | 4.763(1.548-14.653) | 5.138(1.642-16.078) | 0.008 |
| Adjusted^a^ HR(95%CI) | 1.000 | 3.264(1.084-9.826) | 4.086(1.344-12.418) | 4.604(1.498-14.146) | 1.599(1.871-15.634) | 0.019 |
| Adjusted^b^ HR(95%CI) | 1.000 | 2.939(0.972-8.89) | 3.597(1.175-11.102) | 3.982(1.287-12.318) | 4.288(1.36-13.52) | 0.056 |
| Middle 1/3 |  |  |  |  |  |  |
| Crude HR(95%CI) | 1.000 | 2.318(0.635-8.466) | 2.24(0.594-8.455) | 2.33(0.616-8.81) | 3.098(0.824-11.649) | 0.378 |
| Adjusted^a^ HR(95%CI) | 1.000 | 2.278(0.615-8.439) | 2.392(0.627-9.124) | 2.341(0.61-8.994) | 4.096(1.068-15.716) | 0.077 |
| Adjusted^b^ HR(95%CI) | 1.000 | 2.552(0.648-10.052) | 2.886(0.701-11.872) | 2.472(0.596-10.261) | 5.288(1.268-22.054) | 0.022 |
| Lower 1/3 |  |  |  |  |  |  |
| Crude HR(95%CI) | 1.000 | 0.966(0.525-1.78) | 1.147(0.612-2.148) | 1.232(0.61-2.487) | 2.456(1.138-5.299) | 0.023 |
| Adjusted^a^ HR(95%CI) | 1.000 | 0.946(0.508-1.762) | 1.113(0.586-2.111) | 1.287(0.63-2.629) | 2.247(1.016-4.973) | 0.054 |
| Adjusted^b^ HR(95%CI) | 1.000 | 0.939(0.495-1.78) | 1.111(0.573-2.152) | 1.283(0.615-2.674) | 2.034(0.895-4.621) | 0.123 |
